# Venovenous Extracorporeal CO_2_ removal to support ultraprotective ventilation in moderate-severe ARDS: A systematic review and meta-analysis of the literature

**DOI:** 10.1101/2021.10.26.21265546

**Authors:** Elliott Worku, Daniel Brodie, Ryan Ruiyang Ling, Kollengode Ramanathan, Alain Combes, Kiran Shekar

**Affiliations:** Adult Intensive Care Services, the Prince Charles Hospital, Metro North Hospital and Health Service, Brisbane, Queensland, Australia; University of Queensland, Brisbane, Queensland, Australia; Department of Medicine, Columbia University College of Physicians and Surgeons, New York, USA; Center for Acute Respiratory Failure, New York-Presbyterian Hospital, New York, USA; Yong Loo Lin School of Medicine, National University of Singapore; Cardiothoracic Intensive Care Unit, National University Heart Centre, National University Hospital, Singapore; Sorbonne Université, Institute of Cardiometabolism and Nutrition, Paris, France

**Keywords:** acute respiratory distress syndrome (ARDS), extracorporeal carbon dioxide removal (ECCO_2_R), extracorporeal membrane oxygenation (ECMO), ultra-protective lung ventilation (UPV), ventilator-induced lung injury (VILI), driving pressure (ΔP)

## Abstract

**Background:** A strategy that limits tidal volumes and inspiratory pressures, improves outcomes in patients with the acute respiratory distress syndrome (ARDS). Extracorporeal carbon dioxide removal (ECCO_2_R) may facilitate ultra-protective ventilation. We conducted a systematic review and meta-analysis to evaluate the efficacy and safety of venovenous ECCO_2_R in supporting ultra-protective ventilation in moderate-to-severe ARDS.

**Methods:** MEDLINE and EMBASE were interrogated for studies (2000-2021) reporting venovenous ECCO_2_R use in patients with moderate-to-severe ARDS. Studies reporting ≥10 adult patients in English language journals were included. Ventilatory parameters after 24 hours of initiating ECCO_2_R, device characteristics, and safety outcomes were collected. The primary outcome measure was the change in driving pressure at 24 hours of ECCO_2_R therapy in relation to baseline. Secondary outcomes included change in tidal volume, gas exchange, and safety data.

**Results:** Ten studies reporting 421 patients (PaO_2_:FiO_2_ 141.03mmHg) were included. Extracorporeal blood flow rates ranged from 0.35-1.5 L/min. Random effects modelling indicated a 3.56 cmH_2_O reduction (95%-CI: 3.22-3.91) in driving pressure from baseline (p<0.001) and a 1.89 ml/kg (95%-CI: 1.75-2.02, p<0.001) reduction in tidal volume. Oxygenation, respiratory rate and PEEP remained unchanged. No significant interactions between driving pressure reduction and baseline driving pressure, partial pressure of arterial carbon dioxide or PaO_2_:FiO_2_ ratio were identified in metaregression analysis. Bleeding and haemolysis were the commonest complications of therapy.

**Conclusions:** Venovenous ECCO_2_R permitted significant reductions in ΔP in patients with moderate-to-severe ARDS. Heterogeneity amongst studies and devices, a paucity of randomised controlled trials, and variable safety reporting calls for standardisation of outcome reporting.

Prospective evaluation of optimal device operation and anticoagulation in high quality studies is required before further recommendations can be made.

**Key Messages:** *What is the Key Question?:*

- In adult patients with moderate-to-severe acute respiratory distress syndrome (ARDS), can venovenous extracorporeal carbon dioxide removal (ECCO_2_R) support ultraprotective lung ventilation beyond the current standard for protective ventilation in ARDS?

*What is the bottom line?:*

- Systematic review of available data on venovenous ECCO_2_R shows that it can reduce driving pressure in ventilated patients with moderate-to-severe ARDS, supporting ultraprotective ventilation. Prospective measurement of mechanical power, and greater emphasis on safety and patient-centred outcomes is needed.

*Why read on?:*

- This is the first systematic review to exclusively address venovenous ECCO_2_R use in the moderate-to-severe ARDS cohort. We report the degree of lung protection achieved with venovenous ECCO_2_R devices, along with factors potentially limiting widespread adoption.

## Introduction

Acute respiratory distress syndrome (ARDS) accounts for 10% of all intensive care unit (ICU) admissions, yet remains underappreciated by clinicians.^1^ Ventilator-induced lung injury (VILI) potentiates multiorgan dysfunction^2^ and mediates poor outcomes in ARDS. Limiting tidal volumes and airway pressures in patients with ARDS, demonstrated a survival benefit in the seminal ARDS Network trial,^3^ with potentially protective effects when preemptively applied.^4^ To better protect patients with ARDS, there has been growing interest in ultraprotective ventilation, historically defined by more emphatic reductions in tidal volume compared with standard lung protective ventilation. More recently, driving pressure, the quotient of tidal volume divided by static respiratory system compliance (Vt/Crs)^5^ has been shown to independently predict mortality in secondary analyses of ARDS trials.^5-7,8 9^ Mechanical power further incorporates static and dynamic^10^ determinants of stress and strain (including respiratory frequency) thus providing what may be a more complete evaluation of the injurious potential of ventilation practices.^11 12^ Ultra-protective ventilation is an attractive strategy to mitigate ARDS progression, and is feasible without extracorporeal support; accepting however the risk of severe hypercapnic acidosis, and compensatory increases in respiratory rate which effectively abolish reductions in mechanical power that may be expected from lowered driving pressures.^13^ Furthermore, unchecked hypercapnia may be deleterious through altered alveolar fluid clearance,^14^ conflicting effects on immune function,^15^ and unfavourable increases in pulmonary vascular afterload and right ventricular dysfunction.^16^

Extracorporeal membrane oxygenation (ECMO), and extracorporeal CO_2_ removal (ECCO_2_R) both offer carbon dioxide removal, yet the extent of support provided differs (see supplement). Early iterations of ECCO_2_R employed arteriovenous cannulation, however modern venovenous devices typically utilise centrifugal pumps, and in contrast to ECMO, may be performed with smaller, single site access.^17 18^ As the intention of ECMO is typically oxygenation, a large proportion of native cardiac output should be ‘captured’ by the membrane lung^19^ in order to support significant reductions in mechanical power^20,21^ (typically≥3l.min^-1^),^22^ but the relative solubility of carbon dioxide permits very low blood flows (e.g <500ml.min^-1^) to provide meaningful clearance.^17 23 24^ The extent to which ECCO_2_R may facilitate ultra-protective ventilation is contingent on the interplay between the subject’s metabolism, native lung gas exchange, membrane lung efficiency, sweep gas flow rate, and extracorporeal blood flow rate.^25 26^

Uptake and experience with venovenous ECCO_2_R in ARDS is limited^27^, largely due to safety concerns and unclear efficacy.^28^ We undertook a systematic review and meta-analysis of venovenous ECCO_2_R studies in moderate-to-severe ARDS; to evaluate the evidence for efficacy with respect to supporting ultra-protective ventilation as well as the occurrence of complications.

## Methods

This systematic review was conducted and reported in accordance with the Preferred Reporting Items for Systematic Reviews and Meta-Analyses (PRISMA) statement.^29^ The protocol was registered in the PROSPERO database (CRD42020166051); the initial registration accounted for a systematic review only, but was subsequently modified (by consensus amongst authors) in line with emerging definitions of ultraprotective ventilation, and with the publication of major studies in this area permitting a meta-analysis to be performed.

### Search Strategy and study selection

We interrogated MEDLINE and EMBASE databases for articles from the 1^st^ January, 2000, until 1^st^ September, 2021, using Medical Subject Headings (MeSH) terms and keywords (supplement), adapted from a recent review.^30^ This epoch was justified by the scarcity of pre-2000 evidence, and the advances in general ICU care and extracorporeal support in the last 20 years. A similar time period was selected in a recent meta-analysis pertaining to the use of ECMO.^31^ Citations were screened at the title and abstract level; those of relevance were reviewed in detail. Reference lists were explored, and periodic review of the search was performed. Articles in English reporting on the use of venovenous ECCO_2_R with blood flow rate (BFR) <2.0L.min^-1^ for at least 24 hours in ≥10 mechanically ventilated adult patients with baseline partial pressure of arterial oxygen to fraction of inspired oxygen (PaO_2_:FiO_2_) ratio ≤200 mmHg were considered; American-European Consensus Conference (AECC),^32^ and Berlin^33^ definitions for ARDS were accepted. Abstracts or poster citations, pre-clinical, animal, paediatric and review articles were excluded. Studies utilising pumpless arterial ECCO_2_R devices were also excluded. Authors were contacted for additional clarification where necessary, the absence of which led to study exclusion (figure 1).

**Figure 1a.**
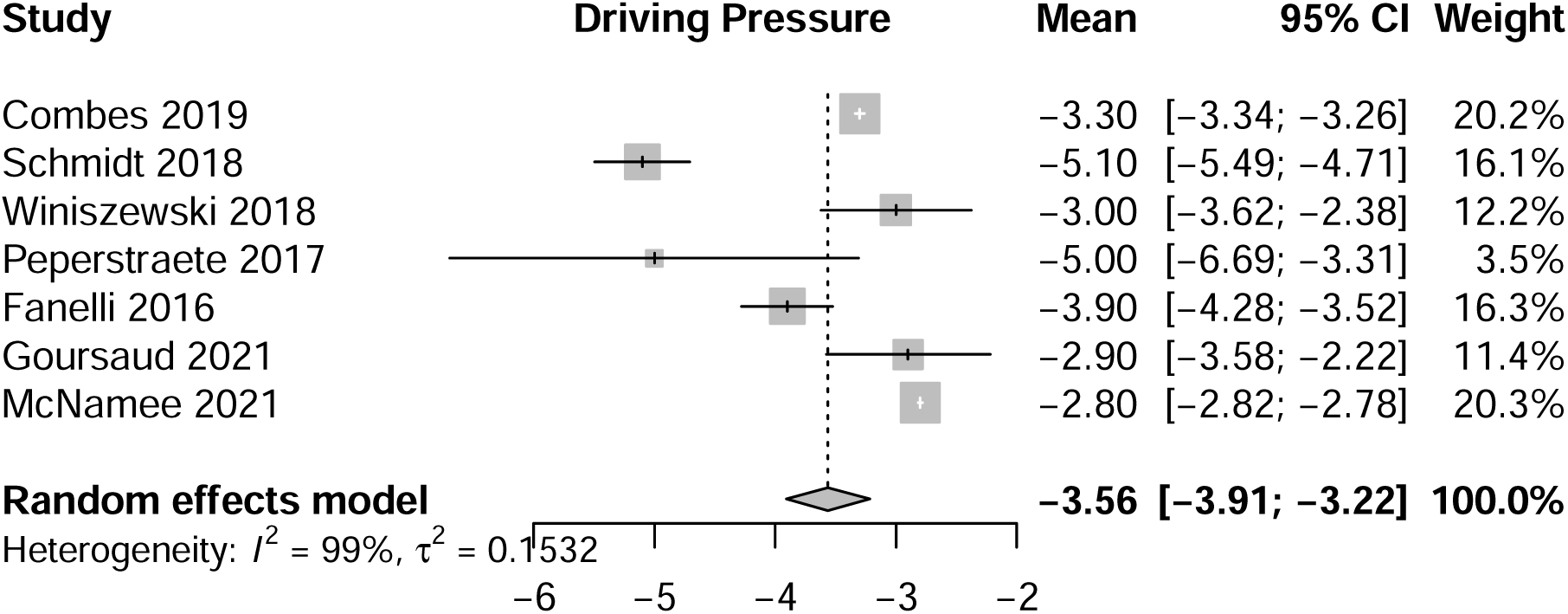
Forest Plot: Change in Driving Pressure following 24hours of ECCO_2_R

**Figure 1b.**
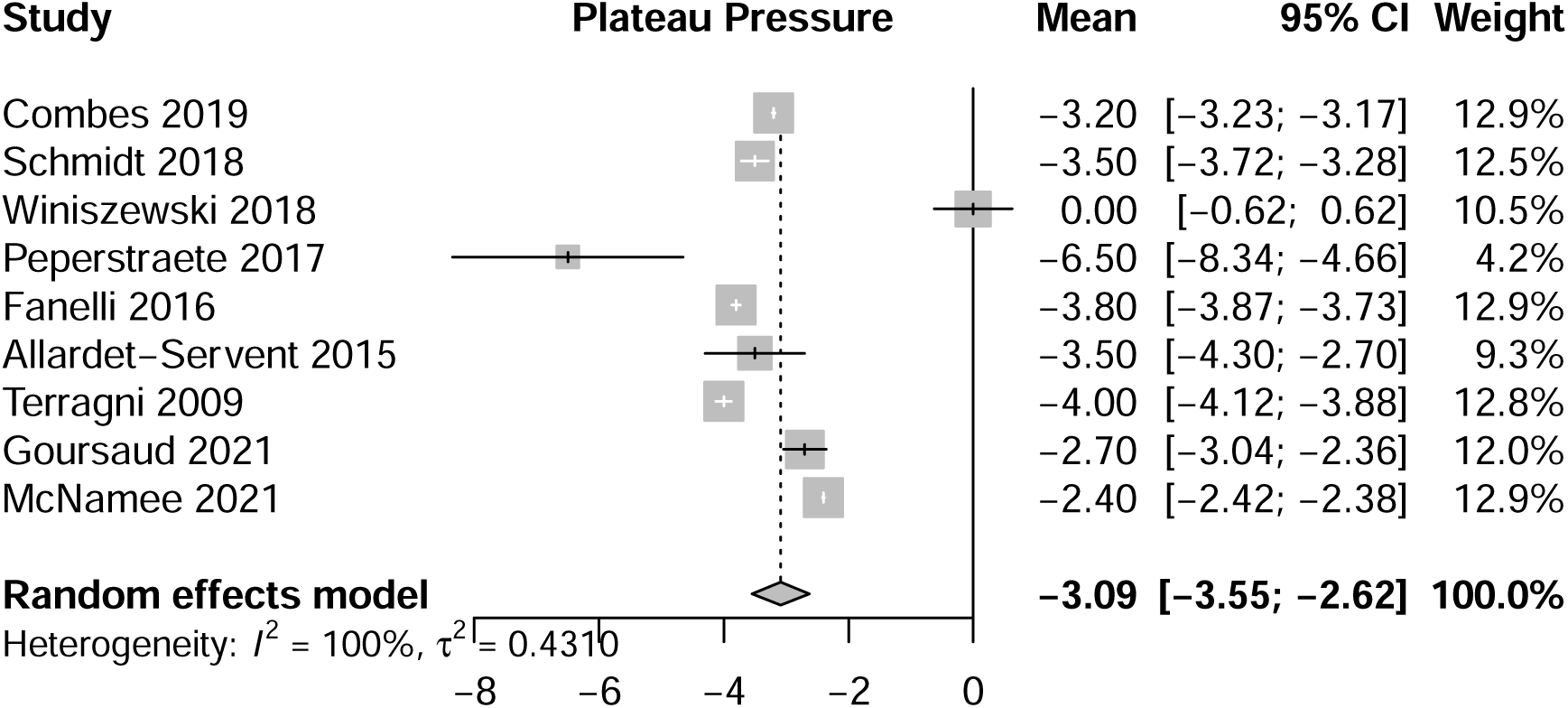
Forest Plot: Change in Plateau Pressure following 24hrs of ECCO_2_R

**Figure 1c.**
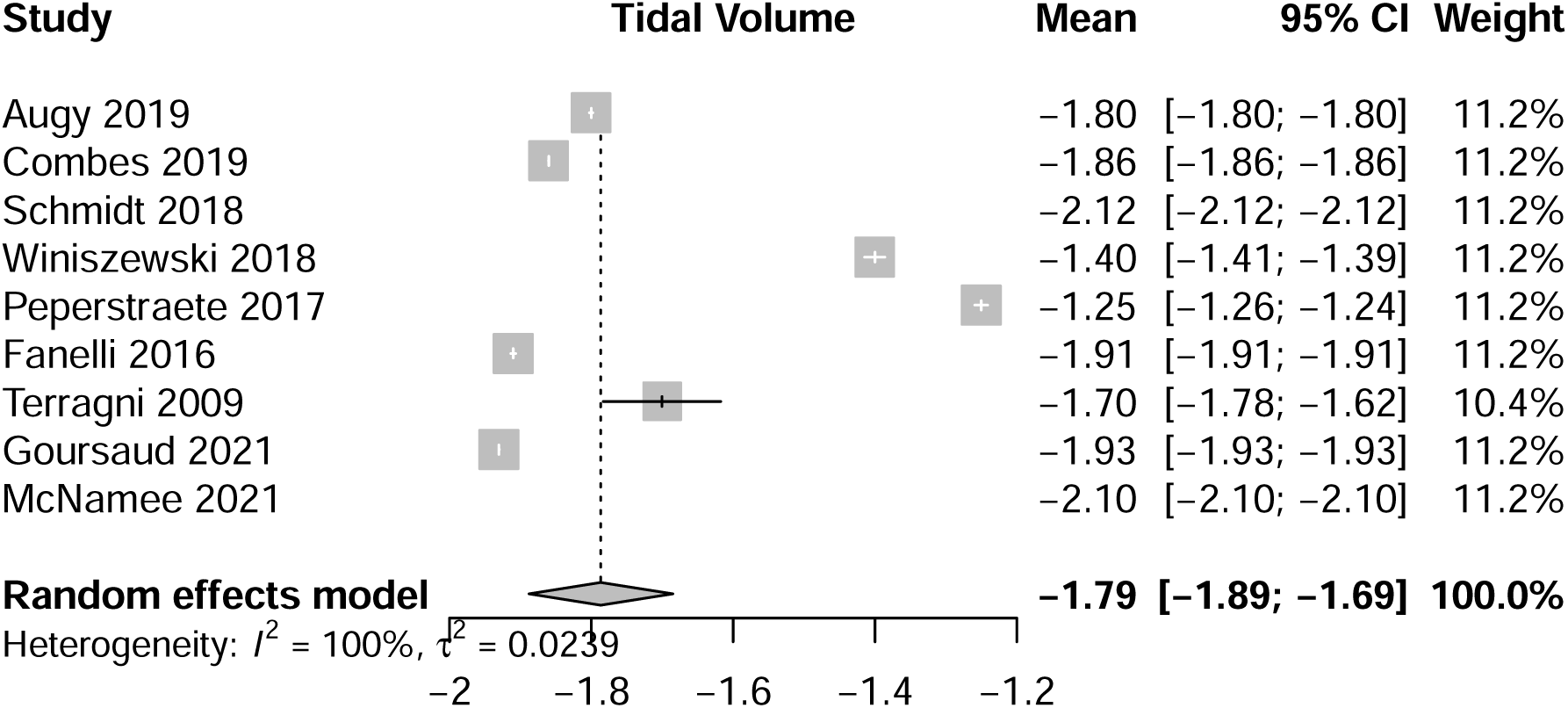
Forest Plot: Change in Tidal volume following 24hours of ECCO_2_R

**Figure 2a.**
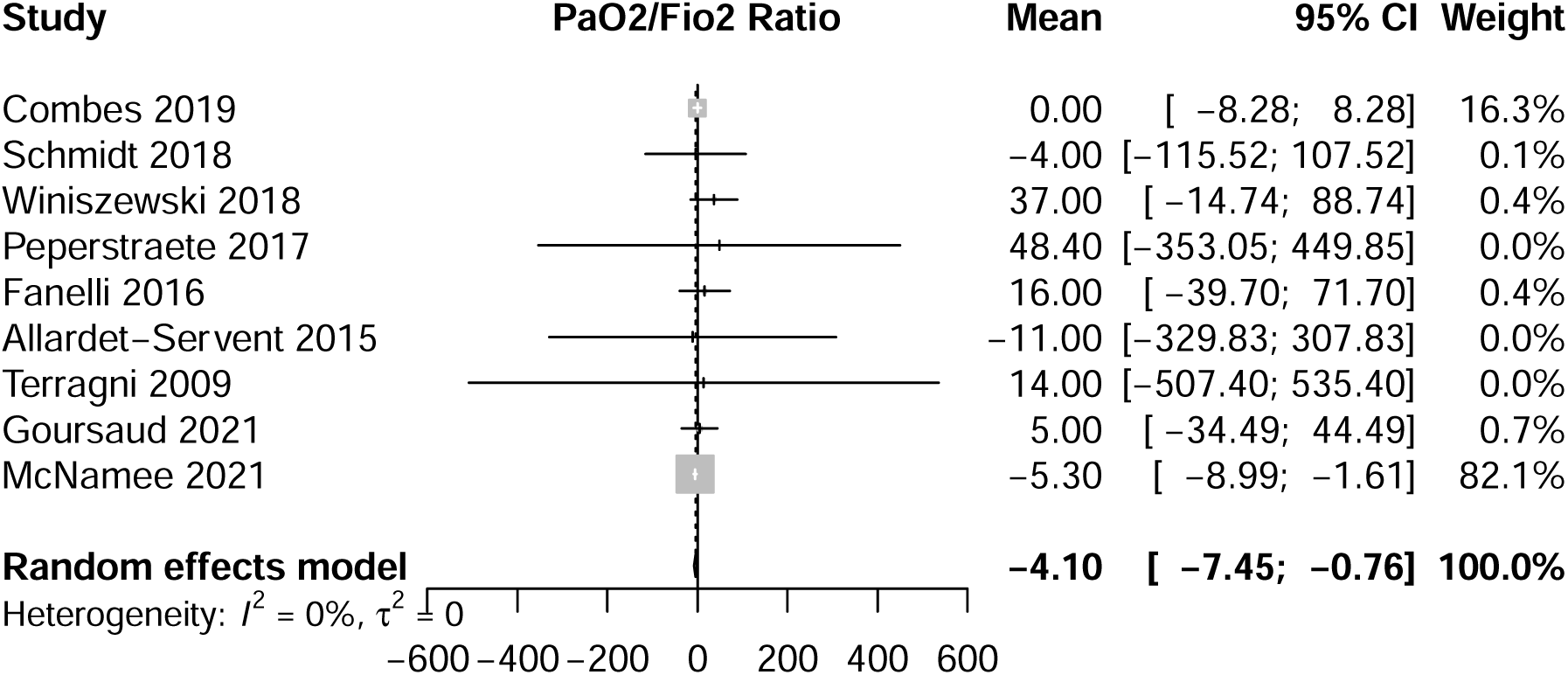
Forest Plot: Change in PaO_2_:FiO_2_ ratio at 24hours of ECCO_2_R

**Figure2b.**
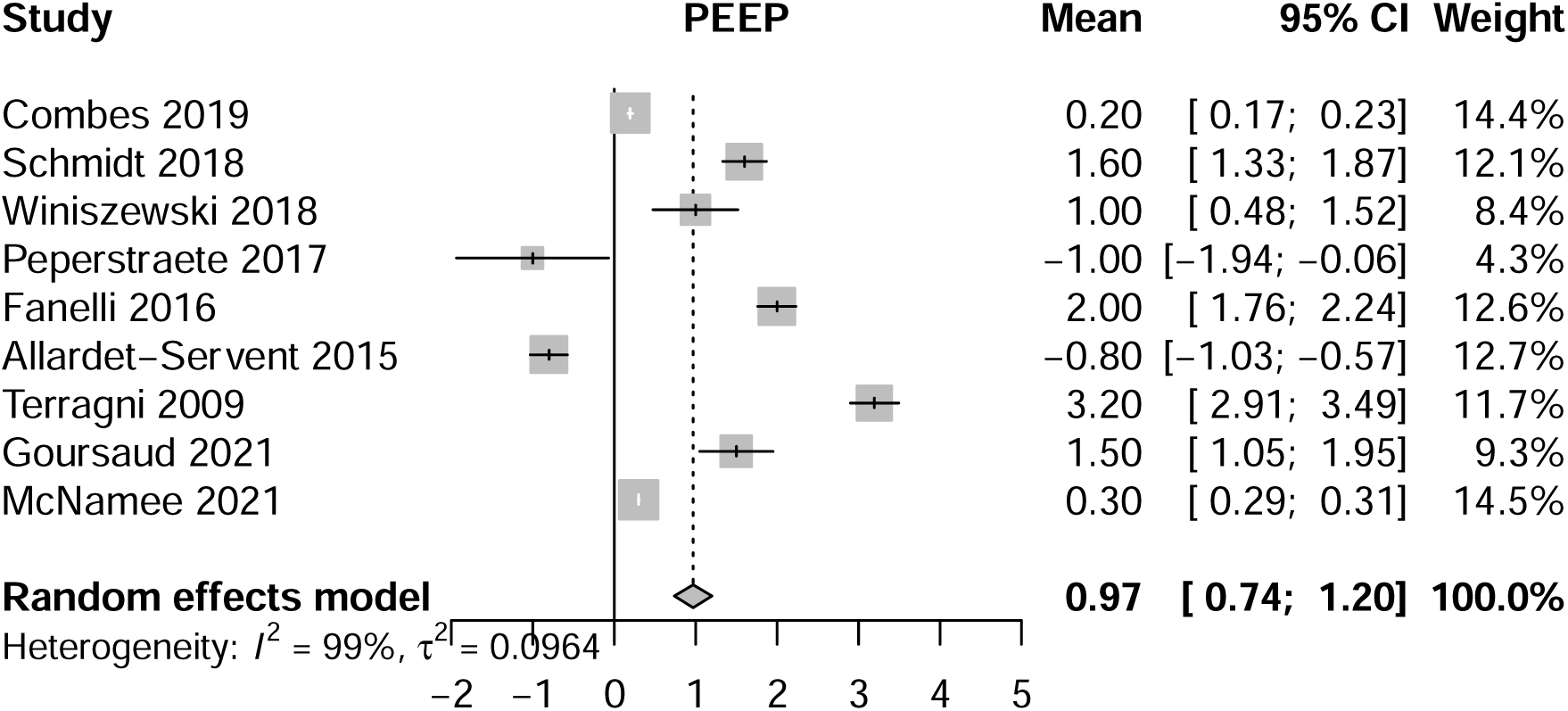
Forest Plot: Change in Positive End Expiratory Pressure at 24hours of ECCO_2_R

**Figure 3a.**
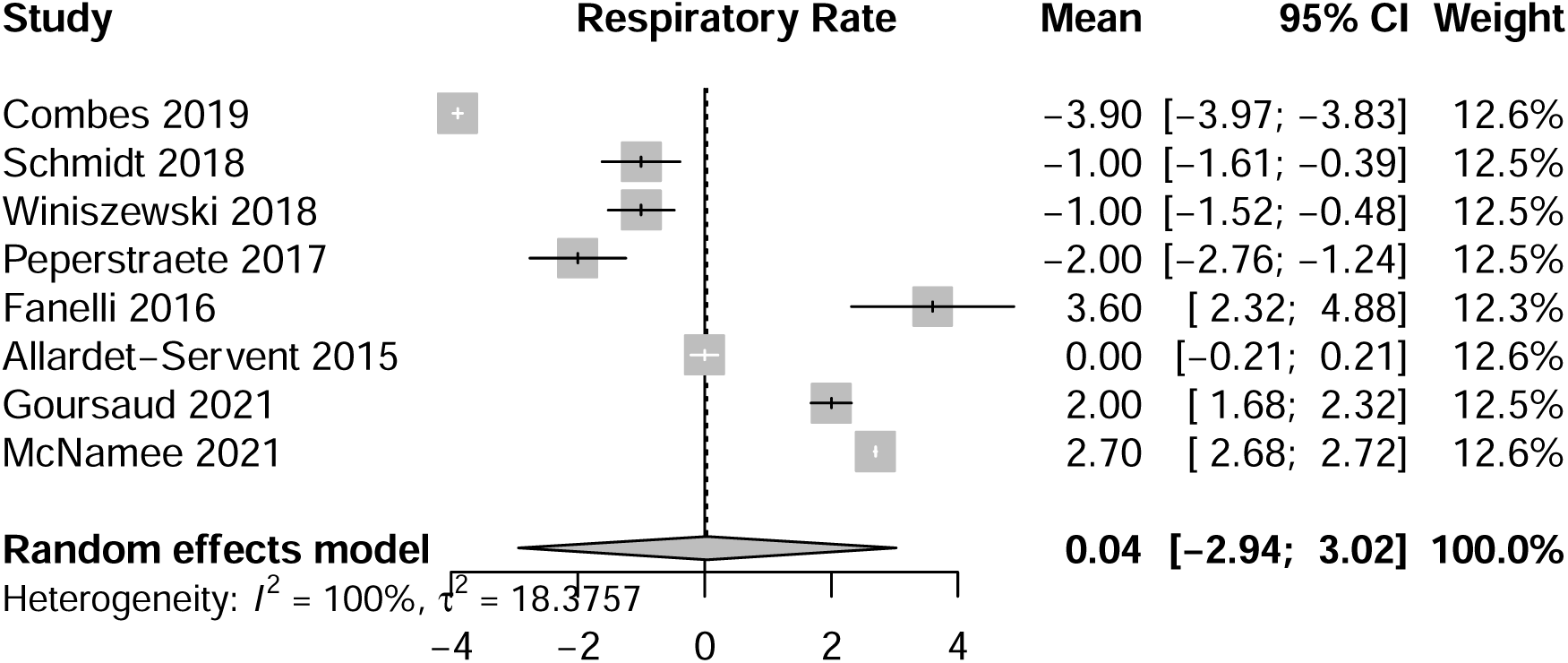
Forest Plot: Change in Respiratory Frequency at 24hours of ECCO_2_R

**Figure3b.**
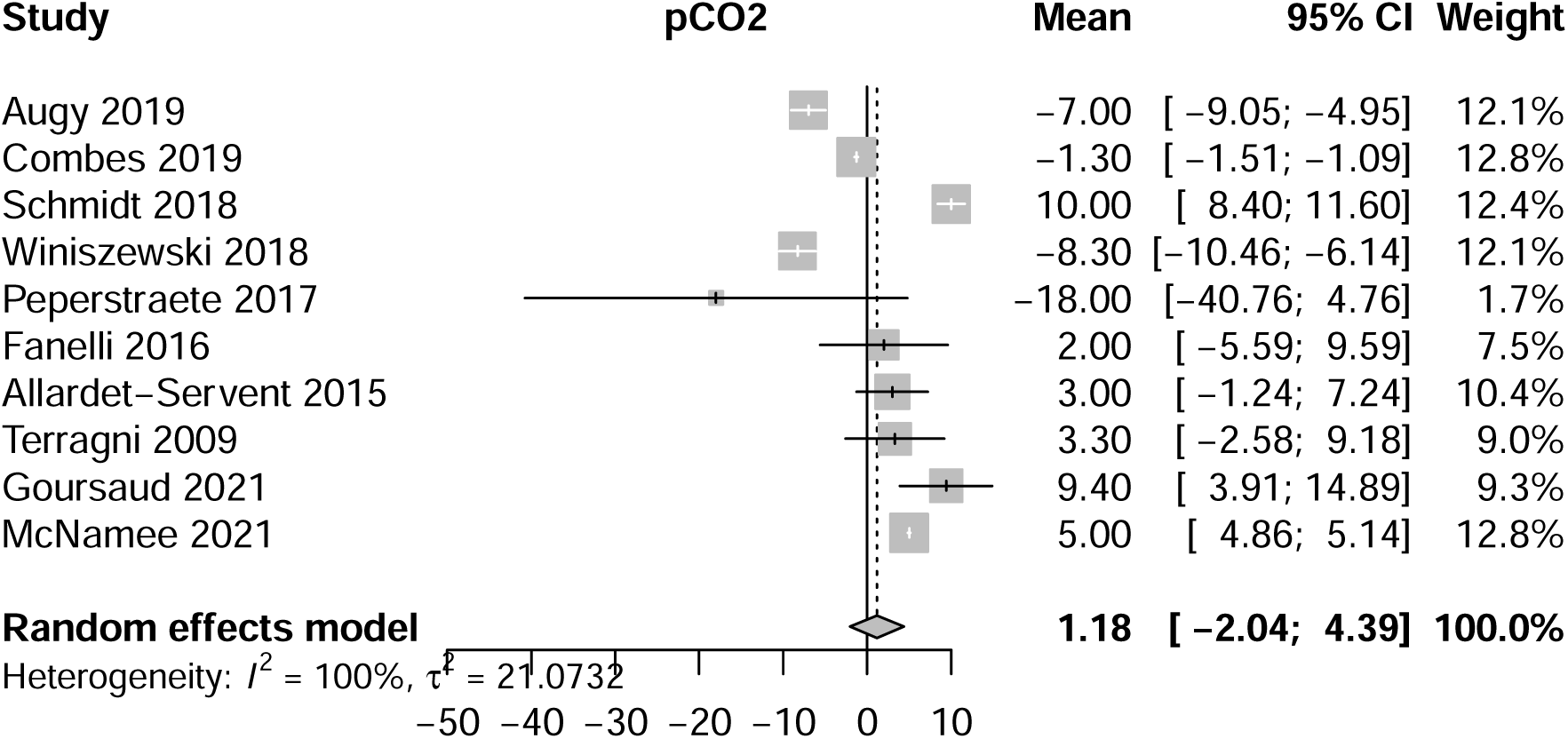
Forest Plot: Change in Arterial Partial pressure of Carbon Dioxide at 24hours of ECCO_2_R

**Figure3c.**
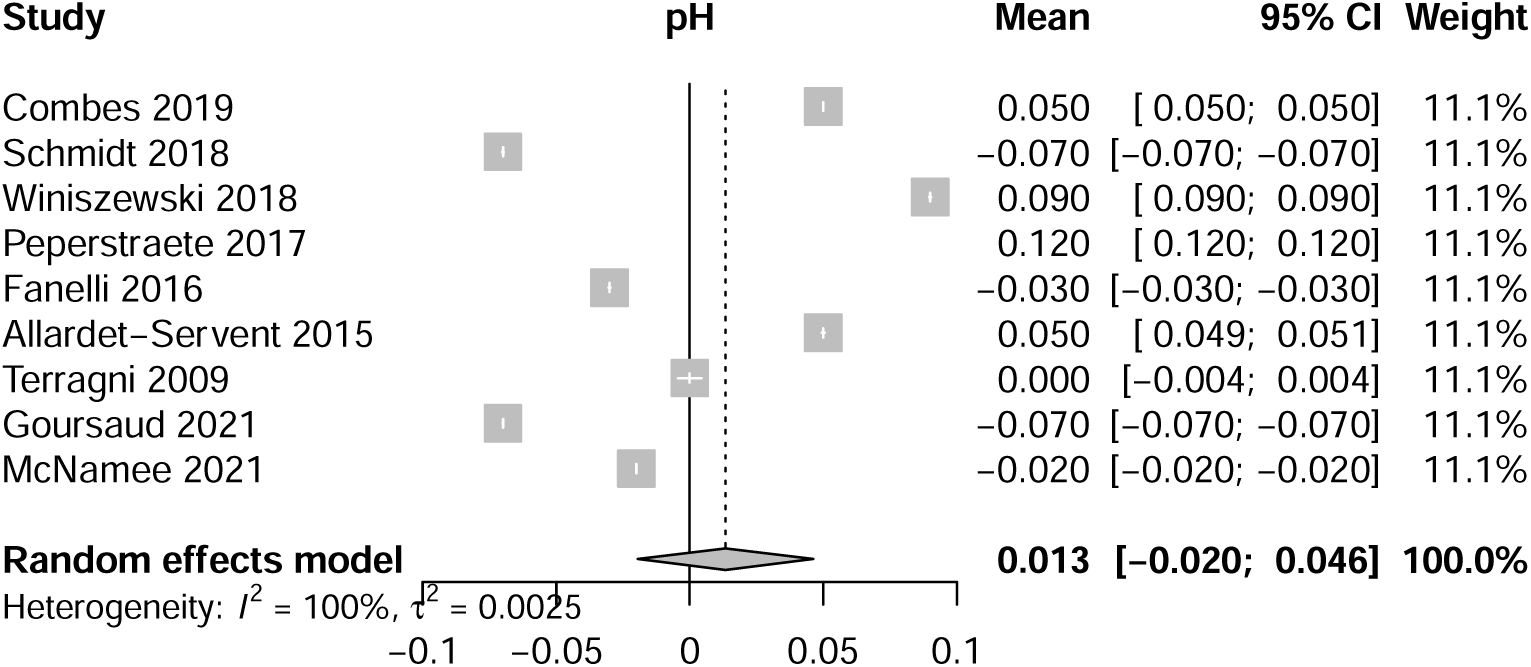
Forest Plot: Change in pH at 24hours of ECCO_2_R

### Assessment of methodological quality

Study quality was assessed using the appropriate Joanna Briggs Institute (JBI) critical appraisal checklists. The certainty of evidence was rated using the Grading of Recommendations, Assessment, Development, and Evaluations (GRADE) approach.

### Outcomes of interest and data collection

The aim of this review was to characterise the evolution of ventilatory and physiological variables, as well as complication rates. The main outcome measure was the change in driving pressure (ΔP) with respect to baseline values; and secondarily the tidal volume (Vt), plateau airway pressure (Pplat), respiratory rate (RR) and positive end-expiratory pressure (PEEP) after 24 hours of venovenous ECCO_2_R therapy. The 24 hour interval was consistently reported across included studies, and chosen as biphasic elimination of CO_2_ under ECCO_2_R has previously been described.^34-37^ Rapid initial clearance is due to removal of the dissolved component, with steady state carbon dioxide removal achieved at approximately 24 hours,^34^ reflecting elimination of stored CO_2_. Data pertaining to study design, patient demographics, and outcome data was extracted to populate tables, designed *a priori* (see supplement). For three studies, graphical data were abstracted using WebPlotDigitiser V4.2.^38^ Data from each study at baseline, and following 24 hours of ECCO_2_R has been summarised in table 3. The mean and standard deviation (SD) were estimated from the median (interquartile range, IQR), as appropriate. In the absence of marked skewness (i.e. the median was approximately midway in the IQR) the mean was estimated as the median and the SD was estimated as IQR/1.35.^39^

### Data synthesis

The primary meta-analysis consisted of pooling the change in ventilatory parameters, while accounting for a pre- and post-intervention correlation by assuming a correlation coefficient (r) of 0.5. We carried out random effects meta-analysis (DerSimonian and Laird, Table 4) based on the Freeman-Tukey double arcsine transformation, and computed the 95% confidence intervals (CIs) using the Clopper-Pearson method.^40-42^ Sensitivity analyses were conducted by simulating different values of r, including 0, 0.25, 0.75 and 0.9. A second sensitivity analysis was conducted by excluding studies with relatively higher risks of bias (defined *a priori* as JBI score <8). As interstudy heterogeneity among observational studies can be misleadingly overestimated when using I^2^ statistics, we used the GRADE approach to assess the interstudy variability^43^. Meta-regression analyses were conducted if a minimum of 6 data points could be collected, in accordance with previous meta-analyses,^43^ to explore potentially prognostically relevant study-level covariates and possible sources of heterogeneity (supplement), for example the influence of baseline driving pressure on the primary outcome under ECCO_2_R. Statistical analyses were performed on R3.6.2.

## Results

Of 534 citations identified, 346 were reviewed at the abstract level, and 23 as full texts. In total, ten studies, reporting 421 patients, were identified spanning October 2009^44^ through August 2021^45^ (PRISMA figure – supplement). There were 2 prospective pilot studies, 4 prospective observational cohort studies, 1 retrospective chart review, 1 prospective multicentre international phase 2 study, 1 quasi-experimental prospective observational cohort study, and 1 randomised controlled trial (RCT) (table 1). Baseline patient demographraphics were broadly homogenous (table1, and supplement) with a preponderance of men (68.4%), and mean age 62.2 years. Baseline PaO_2_:FiO_2_ ratios ranged from 83-173mmHg; two studies^46 47^ included mixed indications for ECCO_2_R therapy (e.g. acute hypercapnic exacerbations of chronic obstructive disease), but ARDS specific data was able to be extracted. In one study, mean Vt at inclusion was slightly above the average at 6.9ml/Kg PBW and was reduced more modestly to <6ml/Kg^48^, with the primary intent of maintaining PaCO_2 (_Table 2). All studies reported pumped venovenous devices, predominantly coupled with double-lumen catheters inserted into the internal jugular vein. Venous access generally ranged from 13Fr-15Fr in size, with larger 24Fr catheters coupled with higher blood flow rates in one trial^49^. Mean extracorporeal blood flows ranged from 350 ml.min-1 to a maximum of 970 ml.min-1 (supplement). Further appraisal of commercially available ECCO_2_R devices, qualities and operational characteristics is provided in the supplement.

**Table 1.**
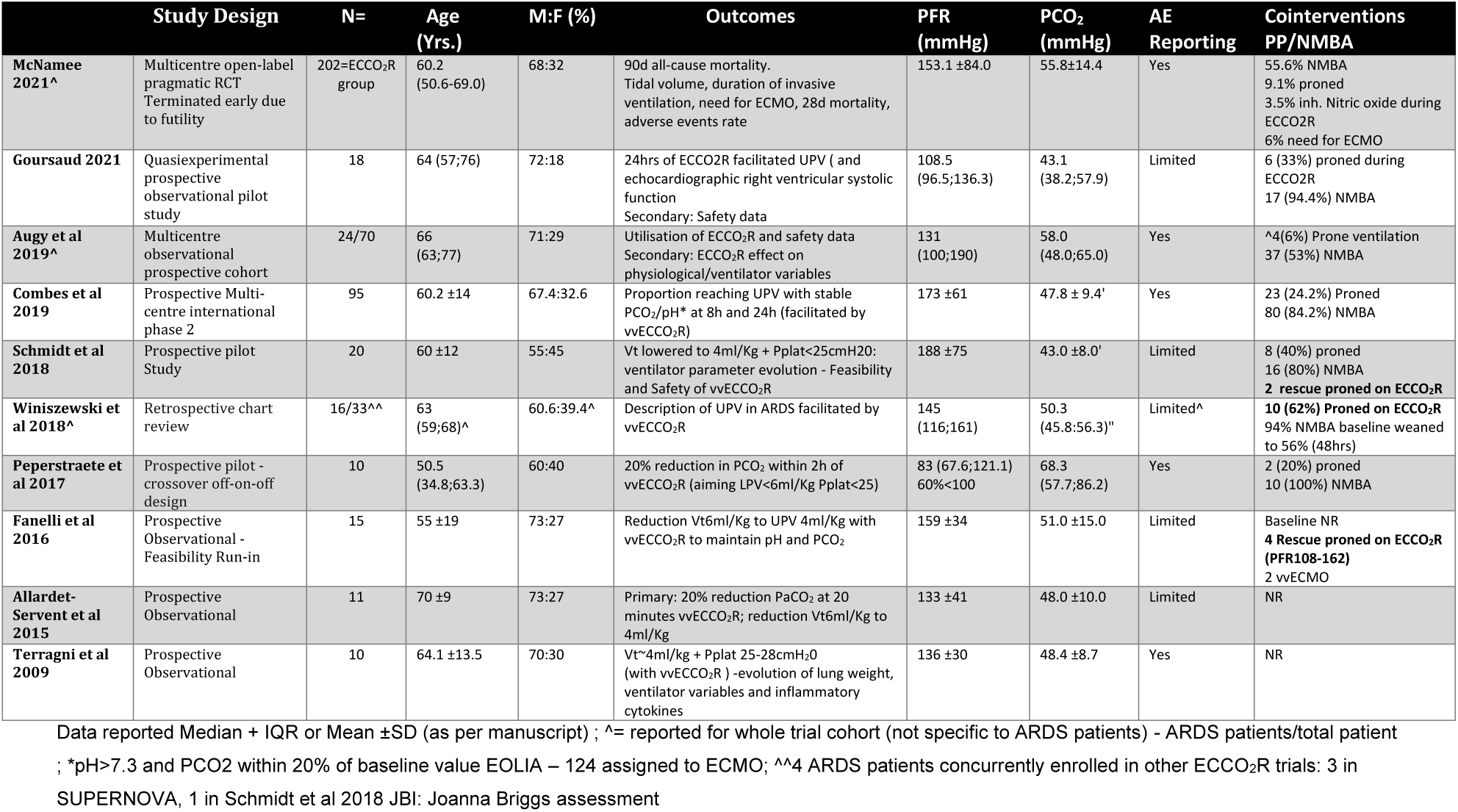
Included studies and demography.

**Table 2.** Ventilator and Physiological Parameters – Baseline Vs 24hrs of ECCO2R *P<0.05 Vs Baseline – reported in source manuscripts NR=Not reported Terragni 2009 was abstracted from graphically presented data = Mean (Min;Max) other trials report as Mean±SD or Median (IQR)

|  | Baseline | 24hrs ECCO2R | Baseline | 24hrs ECCO2R | Baseline | 24hrs ECCO2R | Baseline | 24hrs ECCO2R |
| --- | --- | --- | --- | --- | --- | --- | --- | --- |
| McNamee 2021 | 6.3 (5.8-7.0) | 4.2 ±1.2 | 23.9 ±5.02 | 26.6 ±5.7 | 14.9 ±5.3 | 12.1±4.9 | 25.9±5.2 | 23.5±5.2 |
| Goursaud 2021 | 6.06 (6.00;6.35) | 4.13 (4.0;4.28)* | 28 (25;30) | 30(27.5;32) | 11.5 (10-17.8) | 8.6 (6-11.5) | 25.5(24;28) | 22.5(19.8;25.3)* |
| Augy 2019 | 5.9 (5.5;6.0) | 4.1 (3.9;4.8)* | NR | - | - | - | 28±1.58 | - |
| Combes 2019 | 6.02±0.37 | 4.16±0.46* | 27.4±4.7 | 23.5±6.7* | 13.2±4.1 | 9.9±4.3* | 26.7±3.1 | 23.5±3.9* |
| Schmidt 2018 | 6.10±0.3 | 3.98±0.18* | 26±4 | 25±6 | 13.0±4.8 | 7.9±3.2* | 26.3±3.5 | 22.8±2.6* |
| Winiszewski 2018 | 5.3 (4.4;5.9) | 3.9 (3.5;4.2)* | 26 (22;28) | 21 (18;23)* | 10 (8;15) | 7 (6;10)* | 26 (24;27) | 26 (22;29) |
| Peperstraete 2017 | 6.85 (6.31;7.48) | 5.60(4.90;6.05) | 32.0 (27.8;33.0) | 30.0(27.5;31.5) | 19.0 (17.5;24.0) | 14.0 (11.0;18.5) | 31.5 (28.8;35.5) | 25.0 (22.5;30.5) |
| Fanelli 2016 | 6.2±0.7 | 4.29±0.5* | 28±7 | 31.6±4.6* | 13.8±3.4 | 9.9±3.3* | 27.7±1.6 | 23.9±1.0* |
| Allardet-Servent 2015 | 383±63 (ml) | 269±50 (ml) | 20±2 | 20±2 | - | - | 25±4 | 21.5±3.7* |
| Terragni 2009 | 6.3±0.2 | 4.6(3.6;5.4)* | 31.2±2.3 | - | - | - | 29.1±1.2 | 25.1(23.9;26.0)* |
|  | PaO<sub>2</sub>:FiO<sub>2</sub> ratio |  | PEEP |  | PaCO<sub>2</sub> |  | pH |  |
|  | Baseline | 24hrs ECCO2R | Baseline | 24hrs ECCO2R | Baseline | 24hrs ECCO2R | Baseline | 24hrs ECCO2R |
| McNamee 2021 | 153.1±84.0 | 147.8±52.5 | 11.0 ±3.1 | 11.3 ±3.2 | 55.8 ±14.4 | 60.8 ±14.6 | 7.31±0.09 | 7.29±0.13 |
| Goursaud 2021 | 108.5 (96.5;136.3) | 113.5 (81.9;142) | 11.5 (9.3;14.8) | 13.0 (9.5;15.3) | 43.1 (38.2;57.9) | 52.5 (44.2;64.0)* | 7.38 (7.34;7.42) | 7.31 (7.27;7.33) |
| Augy2019 | 131 (100;190) | - | 10 (5;15.5) | - | 58.0 (48.0;65.0) | 51.0 (44.5;55.7) | 7.24 | 7.31 (7.24;7.36)* |
| Combes 2019 | 168±64 | 168±62 | 13.6±3.7 | 13.8±3.9 | 48.0±9.6 | 46.7±10.4 | 7.34±0.09 | 7.39±0.08* |
| Schmidt 2018 | 188±75 | 184±67 | 13.4±3.6 | 15.0±3.4 | 43±8 | 53±9* | 7.39±0.1 | 7.32±0.1* |
| Winiszewski 2018 | 145 (116;161) | 182(149;211) | 13 (10;15) | 14 (12;18) | 50.3 (45.8;56.3) | 42.0 (36.0;57.0) | 7.31 (7.25;7.41) | 7.40 (7.33;7.45) |
| Peperstraete 2017 | 83.0 (67.6;105.4) | 131.4 (90.2;212.7) | 12.0 (9.0;12.75) | 11.0 (8.5;14.5) | 68.3 (57.3;86.2) | 50.3 (42.9;63.9) | 7.21 (7.11;7.23) | 7.33 (7.29;7.38) |
| Fanelli 2016 | 159±34 | 175±45 | 12±3 | 14±2 | 51±15 | 53±15 | 7.36±0.1 | 7.33±0.1 |
| Allardet-Servent 2015 | 150 (97;201) | 139(87;191) | 11.7±2.2 | 10.9±1.9 | 47±10 | 50±7 | 7.28±0.12 | 7.33±0.08* |
| Terragni 2009 | 135±30 | 149 (109;248) | 12.1±2.5 | 15.3(14.0;16.0)* | 48.4±8.7 | 51.7 (43.8;58.0) | 7.36±0.03 | 7.36(7.30;7.41) |

**Table 3a.**
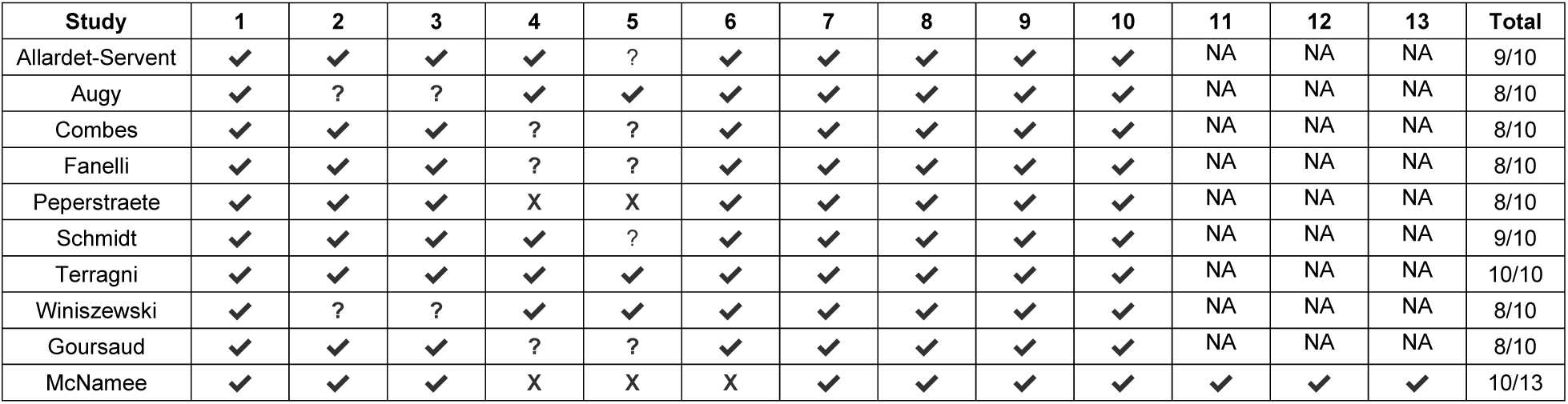
Joanna Briggs Institute Critical Appraisal Checklists for case series and randomised controlled trials (McNamee)

**Table 3b.**
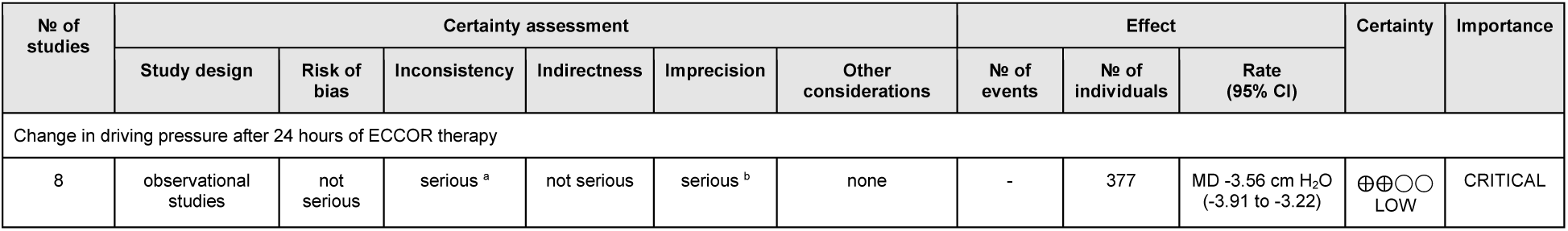
Grading of Recommendations, Assessment, Development, and Evaluations Explanations a. There was significant heterogeneity, and visual inspection of the forest plots found that the point estimates were rather sparsely distributed; confidence intervals occasionally overlapped. b. While the 95%-CI remains relatively narrow with respect to the pooled estimate, the sample size is small and below the optimal information size. As such, we downgraded for imprecision.

The results of the meta-analysis assuming an r-value of 0.5 are presented here; the results of the other analyses with different r-values can be found in table (4). With random effects modelling, the pooled reduction in ΔP was 3.56 cmH_2_O (95%CI: -3.91 to -3.22, table 4, low certainty). The pooled estimates for the remaining variables, revealed a reduction in Vt of –1.79ml/Kg (95%CI: -1.89 to -1.69) and a correspondent change in Pplat of –3.09 cmH_2_O (95%CI: -3.55 to -2.62). Under ECCO_2_R, the PaO_2_:FiO_2_ ratio was slightly reduced by 4.1mmHg (95%CI: -7.45 to -0.76), while PEEP was increased by 0.97cmH_2_0 (95%CI: 0.74-1.20). Forest plots for ΔP and Vt can be found in figures 4 and 5. Confidence intervals for all remaining variables were nonsignificant and crossed zero; the corresponding forest plots are presented in the supplement. JBI appraisal of the included studies found all studies to be of high quality (JBI score ≥8), precluding any sensitivity analysis based on the risk of bias (table 3a). Meta-regression analysis found that increasing age was associated with smaller reductions in ΔP (regression coefficient: 0.13, 95%CI: 0.02 to 0.24, p=0.020), but there were no other significant interactions between demographics, baseline ΔP, PaCO_2_, and PaO_2_:FiO_2_ ratio on the primary outcome of ECCO_2_R-facilitated reductions in ΔP (see supplement).The overall pooled 28-day mortality in the cohort was 41.6% (95%CI 30.0%-53.5%) (supplement).

**Table 4.**
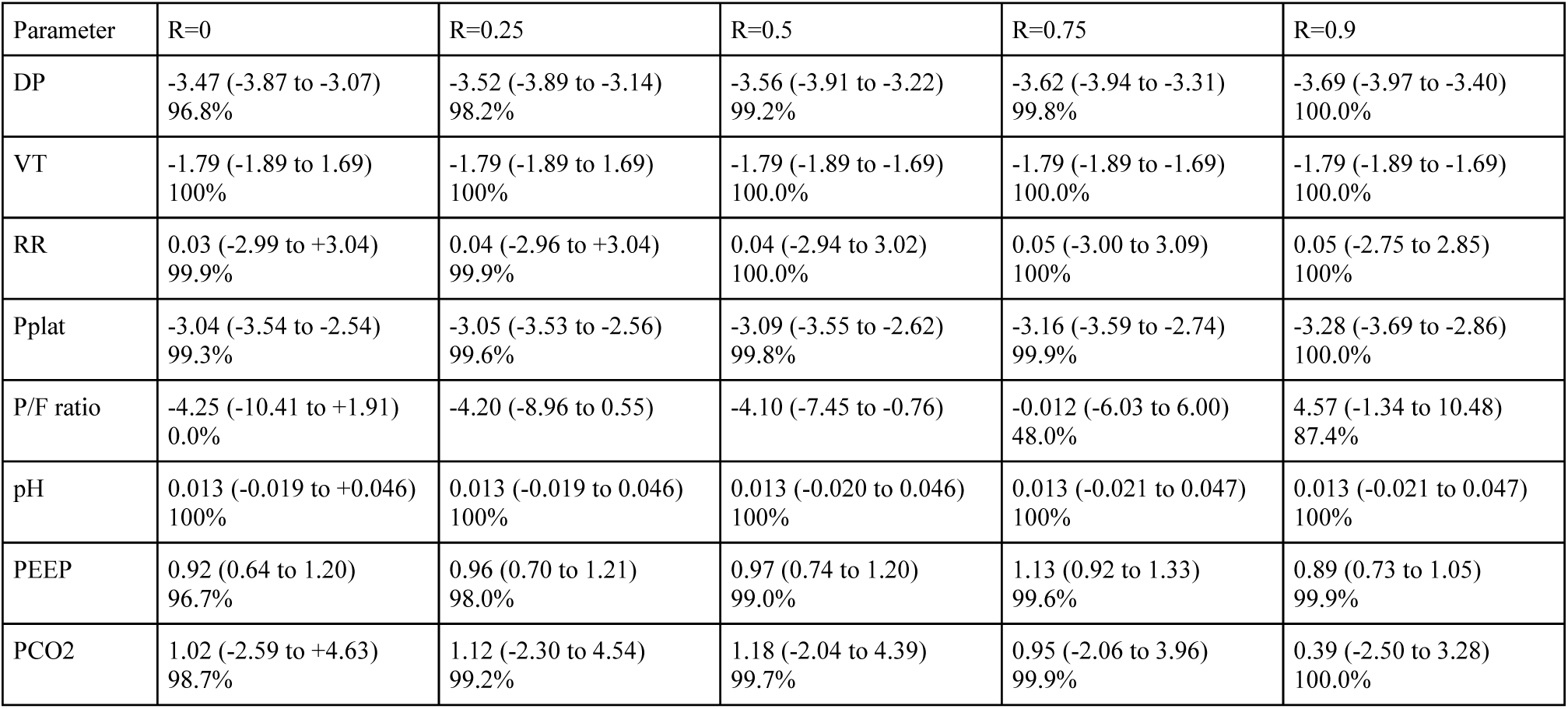
Outcomes of 24hr ECCO2R therapy: results of random effects Meta-analysis

| Parameter | R=0 | R=0.25 | R=0.5 | R=0.75 | R=0.9 |
| --- | --- | --- | --- | --- | --- |
| DP | -3.47 (-3.87 to -3.07)<br>96.8% | -3.52 (-3.89 to -3.14)<br>98.2% | -3.56 (-3.91 to -3.22)<br>99.2% | -3.62 (-3.94 to -3.31)<br>99.8% | -3.69 (-3.97 to -3.40)<br>100.0% |
| VT | -1.79 (-1.89 to 1.69)<br>100% | -1.79 (-1.89 to 1.69)<br>100% | -1.79 (-1.89 to -1.69)<br>100.0% | -1.79 (-1.89 to -1.69)<br>100.0% | -1.79 (-1.89 to -1.69)<br>100.0% |
| RR | 0.03 (-2.99 to +3.04)<br>99.9% | 0.04 (-2.96 to +3.04)<br>99.9% | 0.04 (-2.94 to 3.02)<br>100.0% | 0.05 (-3.00 to 3.09)<br>100% | 0.05 (-2.75 to 2.85)<br>100% |
| Pplat | -3.04 (-3.54 to -2.54)<br>99.3% | -3.05 (-3.53 to -2.56)<br>99.6% | -3.09 (-3.55 to -2.62)<br>99.8% | -3.16 (-3.59 to -2.74)<br>99.9% | -3.28 (-3.69 to -2.86)<br>100.0% |
| P/F ratio | -4.25 (-10.41 to +1.91)<br>0.0% | -4.20 (-8.96 to 0.55) | -4.10 (-7.45 to -0.76) | -0.012 (-6.03 to 6.00)<br>48.0% | 4.57 (-1.34 to 10.48)<br>87.4% |
| pH | 0.013 (-0.019 to +0.046)<br>100% | 0.013 (-0.019 to 0.046)<br>100% | 0.013 (-0.020 to 0.046)<br>100% | 0.013 (-0.021 to 0.047)<br>100% | 0.013 (-0.021 to 0.047)<br>100% |
| PEEP | 0.92 (0.64 to 1.20)<br>96.7% | 0.96 (0.70 to 1.21)<br>98.0% | 0.97 (0.74 to 1.20)<br>99.0% | 1.13 (0.92 to 1.33)<br>99.6% | 0.89 (0.73 to 1.05)<br>99.9% |
| PCO2 | 1.02 (-2.59 to +4.63)<br>98.7% | 1.12 (-2.30 to 4.54)<br>99.2% | 1.18 (-2.04 to 4.39)<br>99.7% | 0.95 (-2.06 to 3.96)<br>99.9% | 0.39 (-2.50 to 3.28)<br>100.0% |

## Complications

The mechanical, and patient-related complications of venovenous ECCO_2_R are summarised in Table 4, with expansion in the supplementary materials. Importantly, not all studies documented all complications. Anticoagulation strategies varied, with 4 studies utilising activated partial thromboplastin time/ratio, 2 studies recording activated clotting time, and 3 adjusting to anti-factor Xa levels; in the most recent study, the intensity of anticoagulation was not reported. Bleeding rates where reported, ranged 10-50% (5/10 studies), with *clinically significant* bleeding rates - variably defined by transfusion requirement and validated tools (see table 4) - between 6 and 50%. Intracranial haemorrhage was reported in 5 studies ^45-47 49 50^ at a rate of between 1 and 5%;. Mechanical complications included catheter dislodgement, kinking or thrombosis ranging 4-40% (6/10 studies), with 2 trials reporting catheter-related infections, n=3.^46 49^ Circuit thrombosis was documented in 7 studies ranging from 14-30%, leading to circuit changes in up to 6% of cases in one series. Air was noted in 2 extracorporeal circuits in one study,^46^ without clinically apparent air embolism.

## Discussion

To our knowledge, this is the first systematic review evaluating venovenous ECCO_2_R in moderate-to-severe ARDS. Two contemporary systematic reviews assessed mixed cohorts of both chronic obstructive pulmonary disease and ARDS patients,^30^ or included pumpless arteriovenous devices;^51^ neither evaluated the larger venovenous ECCO_2_R study published by Combes et al^49^ or the recently published “pRotective veEtilation with venovenouS lung assisT in respiratory failure (REST)” trial^45^. Our main finding was a significant 3.56cmH_2_0 reduction in driving pressure at 24 hours. Ventilatory parameters under ECCO_2_R largely conformed with ultra-protective ventilation targets from a mechanical perspective (driving and plateau pressures) ^52^. Plateau pressure was ≥25cmH_2_0 in all 10 cohorts at the outset, and was reduced significantly at 24 hours with reported values ≤25cmH_2_0 in 7 studies. Driving pressure was >14cmH_2_0 in just two of the seven studies reporting ΔP at baseline. While it was reduced to <10cmH_2_0 in 5 studies, and below 14cmH_2_0 in all reporting studies, tidal volume remained slightly above 4ml/Kg in the majority (table 2). This suggests that studies reported patients with relatively preserved respiratory compliance, in whom ECCO_2_R may not be maximally efficacious.^53 54^

Whether progressive reductions in driving pressure below conventionally accepted limits of ultraprotective targets can further improve patient centred outcomes compared with standard low-volume, low-pressure ventilation strategies^6 7 21 55 56^ is not entirely clear. A recent large RCT comparing conventional low tidal volumes with ECCO_2_R-enabled ultraprotective ventilation found no difference in 28-day mortality, a longer duration of mechanical ventilation and ICU length of stay (LOS) and a 31% rate of serious adverse events in the intervention group^45^. Patients with moderate-to-severe ARDS are a heterogenous cohort with emerging clinical sub-phenotypes,^57 58^ thus the absence of mortality benefit in two recent RCTs is unsurprising.^45 59^ Our analysis showed a pooled 28-day mortality was 41.6% (95%CI: 30.0%-53.5%), which is similar to that reported in a large meta-analysis of mechanically ventilated ARDS patients of comparable severity.^9^ The lack of prospective comparison limits inferences, but the similar mortality rates, may indicate an inability of ECCO_2_R to meaningfully attenuate mechanical power, perhaps through underappreciated influences of respiratory frequency and PEEP.^10 55 60^

Reductions in ventilatory frequency may represent a key target in limiting VILI. Expert opinion suggests that ECCO_2_R ultra-protective ventilation should be used to effect a respiratory rate <25 breaths per minute^52^. Greater reductions in ventilator frequency may have been demonstrated if investigators had specifically targeted greater this, had been more permissive of hypercapnia, or if the devices that were used could either achieve higher blood flow rates or greater CO_2_ removal efficiency. The median pH was >7.3 in nine of ten studies at 24hrs (table 2), however it is pertinent that the median PaCO_2_ at 24hrs remained ≥50mmHg in 7 of the nine included studies, and greater than 55mmHg in one (pH 7.29). The absolute degree of hypercapnia tolerated must not only consider in terms of acid-base, but also right ventricular performance.^50^ Interestingly, reductions in plateau pressure after ECCO_2_R initiation, rather than change in PaCO_2_ were best correlated with improvements in right ventricular afterload.^50^ Baseline PaCO_2_ (a surrogate for the underlying dead space fraction) has previously been suggested to predict patients in whom ECCO_2_R could be most efficacious,^53 54^ however this was not evident in the current metaregression analysis, nor was an interaction with severity of ARDS by oxygenation. As lower flow ECCO_2_R devices only permit partial correction of PaCO_2_, sedation and neuromuscular blocking agents may be needed to play a role in modulating patients’ respiratory drive and ventilator asynchronies.^61-63^

Prone positioning (PP) may be synergistic with ECCO_2_R, through enhanced reductions in driving pressure, ^53 54 64^ CO_2_ elimination^65 66^, and right ventricular unloading^67^.While prone positioning was infrequently employed in the reported ECCO_2_R studies (n=22), it was undertaken, when used, without serious incident (table 1).

A trend towards lower thrombosis and bleeding rates was seen in more recent studies, corroborating existing insights.^51^ Two studies from 2016, and 2017 described 46.7% and 50% bleeding rates respectively, while the five papers from 2018-2021 recorded 10-24.3% bleeding rates. Anti-Xa-guided anticoagulation may offer favourable bleeding profiles,^68^ however, this could not be statistically evaluated here and definitions of bleeding were not standardised. Higher blood flow rate devices were associated with reduced haemorrhage and haemolysis (21% vs 6%, p=0.045)Land (27% vs 6%, p=0.01) respectively in SUPERNOVA,^69^ and the same inverse relationship - bleeding: (30% vs 6%, p=0.04), and haemolysis: (28% vs 0%, p=0.03)^46 49^ were documented by Augy et al.^46^ Rotary pumps may exert increased blood trauma at lower blood flows (particularly<1L.min-1),^70-72^ causing shear stress and loss of high molecular weight von Willebrand factor multimers.^73-76^ Mean blood flow rates may not only influence hematologic complications, but also the degree of lung protection achievable;^25 26^ unfortunately several studies utilised a variety of devices and different blood flow rates, such that metaregression analysis was not appropriate. While anticoagulation management is extrapolated from the ECMO experience, perhaps personalised approaches based on device and patient characteristics, even including regional methods of anticoagulation^77^ may enhance ECCO_2_R safety.^51 78^

### Strength and limitations

Strengths of this review include the comprehensive search, broad criteria including available RCT data, and relevant exclusion criteria. Rigorous quality assessment was performed, deeming studies of high quality and suitable for inclusion. Furthermore, we limited the inclusion of studies to moderate-to-severe ARDS which was undertaken in order to homogenise the study population, and meta-regression analyses were specifically performed to interrogate whether ECCO_2_R efficacy was influenced with respect to baseline PaCO_2_, as a surrogate of dead space fraction,^53 54 66^ although no such relationship was identified. Where possible, duplication of data was avoided, however 4 patients were concurrently enrolled in more than one of the reported studies; a lack of patient-level data precluded their identification and removal.

We nonetheless recognise several limitations of our review. Only 10 studies qualified for analysis, of which, just one was an RCT. Over 75% of the patient data was contributed by just 3 studies, with the largest study responsible for nearly half. In the aforementioned study, 50 centres were involved, thus enrolment may have been as low as 4 patients per centre. The other two manuscripts were from highly experienced ECMO centres, which may have influenced their low mortality rates (supplement). A volume-outcome effect^78^ cannot be discounted. In addition, given that these studies were mostly observational, the effect estimates are open to confounding factors, particularly in the absence of propensity-score or risk-adjustment methods. Some of these include the ventilatory mode employed, right ventricular function, and other adjuvant therapies (e.g proning), with variability potentially accounting for some of the herteogeneity in observed mortalty rates (15-47%). The meta-regression analysis is limited by a small sample size (6 studies minimum), and open to Type II errors.

Further dividing the moderate-to-severe ARDS category into patients with a PaO_2_FiO_2_ ratio above and below 150mmHg,^79 80^ or even more directly, by dead space fraction and compliance,^53 54 81^ may enhance selection. Aside from survival, there is glaring need to prospectively investigate cost effectiveness of therapy,^82^ and other patient-centred outcomes such as length of hospital stay, delirium, return to premorbid function and long-term outcomes, which were inconsistently reported if at all. A suite of therapies combining ECCO_2_R of adequate intensity with prone ventilation (to perhaps enhance both lung and right ventricular protection)^66 67 83^, early spontaneous breathing^84^ and mobilisation may be required to effect a meaningful survival benefit. Even in light of recent evidence, the question of whether venovenous ECCO_2_R has the power to impact mortality in the moderate-to-severe ARDS cohort still remains.

## Conclusion

Venovenous ECCO_2_R supported significant reductions in driving pressure at 24 hours in moderate-to-severe ARDS, with an overall mortality of 41.6%. While early ECCO_2_R may facilitate ultra-protective ventilation and mitigate ARDS progression, the benefits are currently offset by the invasiveness of therapy, and limited CO_2_ removal made possible at low blood flow rates. Significant reductions in respiratory rate, and hence further reductions in mechanical power may not be feasible when using very low blood flow rates, potentially limiting the utility of ECCO_2_R in patients with more severe forms of ARDS, or right ventricular dysfunction. Enrichment of study populations, and reporting data consistently to minimum standards, is critical to meaningfully researching benefits of ECCO_2_R therapy in ARDS.

## Data Availability

All data produced are available within the main text, supplement, or via reasonable request to the authors.

## Acknowledgements

The authors thank Dr Dominic Worku (St George’s Hospital NHS trust, London) for his contribution with the literature review, Dr. Chuen Seng Tan (Saw Swee Hock School of Public Health, National University of Singapore) for the assistance with the final statistical analysis, and Dr. Karen Hay for her efforts in preliminary statistical analyses. Dr Shekar acknowledges research support from Metro North Hospital and Health Service.

## Contributors

EW and KS conceived the study and protocol for the systematic review. EW produced the original draft and coordinated the writing process. EW performed the data collection. RRL conducted the statistical analysis and was assisted by Dr Karen Hay (QIMR Berghorfer) and Dr Chuen Seng Tan. All authors contributed to the interpretation of the results. DB, KR, and RRL contributed significantly to the editing process. AC provided critical commentary and insights to the ongoing revisions, and all authors approved the final manuscript.

## Data sharing statement

The search strategy and further data analysis is available in the supplementary materials

## MAIN TABLES

1. Included Study Characteristics
2. Ventilation and Physiological Parameters
3. Quality assessment
  a. Joanna Briggs Institute (JBI) Critical Appraisal Checklist for Case Series and Randomised Controlled Trials
  b. GRADE – Grading of Recommendations, Assessment, Development and Evaluations
4. Outcomes of 24hr ECCO2R therapy: results of random effects Meta-analysis
5. Forest Plots 1a/b/c = Driving Pressure, Plateau Pressure, Tidal Volume
6. Forest Plots 2a/b = PaO_2_:FiO_2_ Ratio, Positive End Expiratory Pressure
7. Forest Plots 3a/3b/3c = Respiratory Frequency, PaCO_2_, pH
8. Adverse Events

---

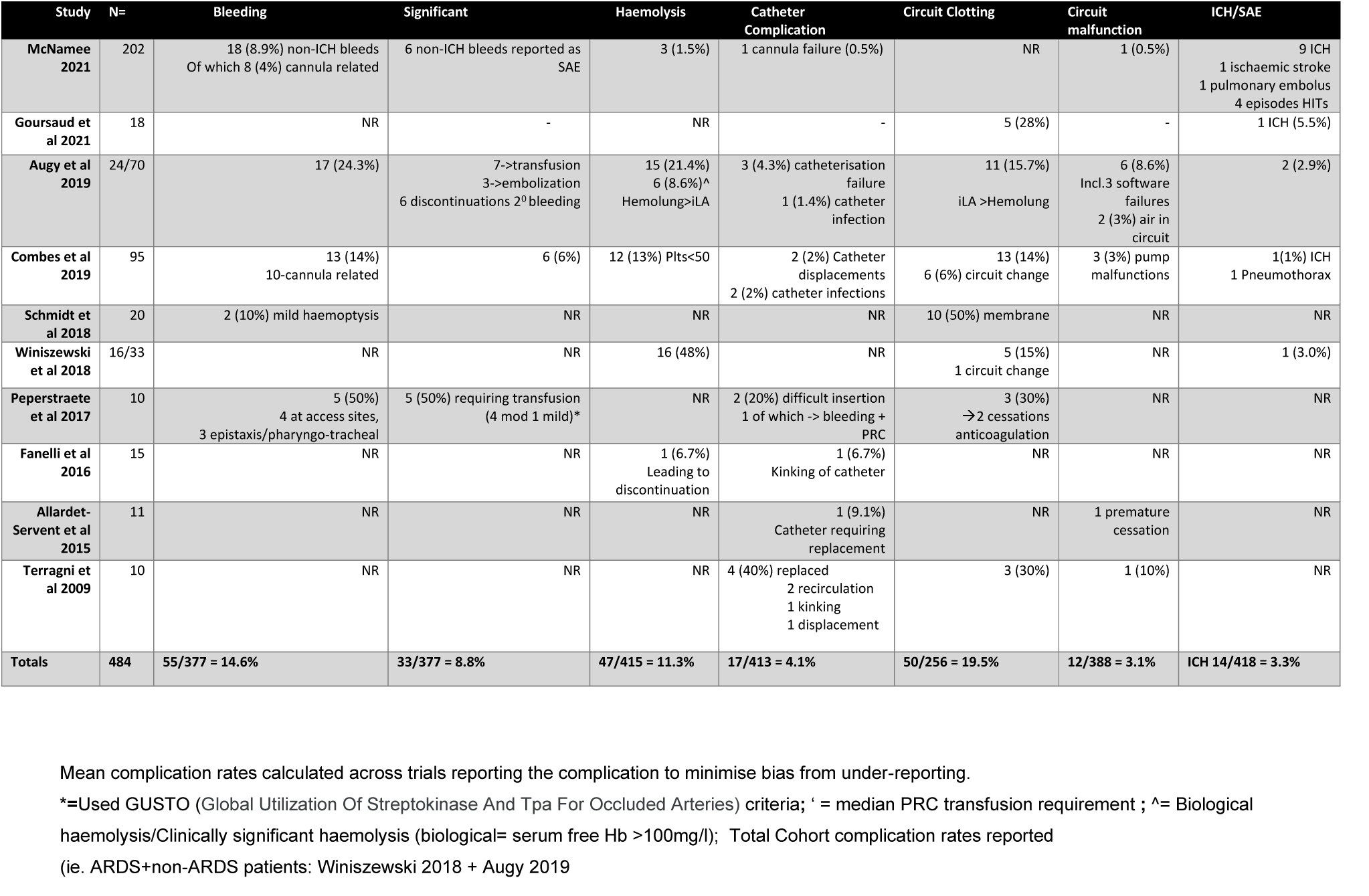

