## Supplementary material for "Venovenous Extracorporeal CO_2_ removal to support ultraprotective ventilation in moderate-severe ARDS: A systematic review and meta-analysis of the literature": 22.10 Final Supplementary (CLEAN).docx

**SUPPLEMENTARY MATERIALS**

- EMBASE/MEDLINE Search Strategy
- Figure 1. PRISMA Flow Diagram
- Table 1.ECCO2R devices used
  - A) By device
  - B) By study
- Table 2. Patient Demographics and Outcome data
- Table 3. Meta-regression Analysis
- Table 4. Bleeding and Anticoagulation

**EMBASE Search Strategy**

(('intensive care unit'/exp OR 'gicu' OR 'gicus' OR 'icu`s' OR 'close attention unit' OR 'combined

medical and surgical icu' OR 'combined surgical and medical icu' OR 'critical care

unit' OR 'general icu' OR 'intensive care department' OR 'intensive care unit' OR 'intensive care

units' OR 'intensive therapy unit' OR 'intensive treatment unit' OR 'medicalsurgery

icu' OR 'medical/surgical icu' OR 'medical/surgical icus' OR 'medico-surgical icu' OR 'mixed

medical and surgical icu' OR 'mixed surgical and medical icu' OR 'respiratory care

unit' OR 'respiratory care units' OR 'special care unit' OR 'surgery/medical icu' OR 'surgicalmedical

icus' OR 'surgical/medical icu' OR 'unit, intensive care') AND ('adult respiratory distress

syndrome'/exp OR 'ards' OR 'acute respiratory distress syndrome' OR 'adult respiratory

distress' OR 'adult respiratory distress syndrome' OR 'lung shock' OR 'posttraumatic lung

failure' OR 'posttraumatic pulmonary insufficiency' OR 'respiratory distress syndrome,

acute' OR 'respiratory distress syndrome, adult' OR 'respiratory distress, adult' OR 'shock lung')

OR 'acute lung injury'/exp OR 'respiratory failure'/exp) AND ('extracorporeal carbon dioxide

removal'/exp OR 'extracorporeal co2 removal' OR 'extracorporeal carbon dioxide

removal' OR ecco2r OR 'extracorporeal carbon dioxide removal device'/exp OR 'low

flow ecmo' OR vvecmo) AND ('controlled study'/exp OR 'randomized controlled trial'/exp OR 'case

control study'/exp OR cohort OR 'feasibility study'/exp OR 'feasibility studies' OR 'feasibility

study' OR 'clinical trial'/exp) AND ('mortality'/exp OR 'carbon dioxide'/exp OR 'pao2 fio2 ratio'/exp

OR 'driving pressure'/exp OR 'tidal volume'/exp OR 'breathing rate'/exp OR 'plateau pressure'/exp)

**MEDLINE Search Strategy**

(((((extracorporeal[All Fields] AND ("carbon dioxide"[MeSH Terms] OR ("carbon"[All Fields] AND

"dioxide"[All Fields]) OR "carbon dioxide"[All Fields]) AND removal[All Fields]) OR ECCO2R[All Fields])

OR ("extracorporeal membrane oxygenation"[MeSH Terms] OR ("extracorporeal"[All Fields] AND

"membrane"[All Fields] AND "oxygenation"[All Fields]) OR "extracorporeal membrane

oxygenation"[All Fields] OR ("extracorporeal"[All Fields] AND "life"[All Fields] AND "support"[All

Fields]) OR "extracorporeal life support"[All Fields])) AND CO2[All Fields]) OR ("extracorporeal

membrane oxygenation"[MeSH Terms] OR ("extracorporeal"[All Fields] AND "membrane"[All Fields]

AND "oxygenation"[All Fields]) OR "extracorporeal membrane oxygenation"[All Fields])) AND CO2[All Fields]

534 potentially eligible articles identified by searches of MEDLINE and EMBASE

346 articles identified for screening at abstract level

24 full manuscripts reviewed

322 articles excluded as

non-English text, animal, paediatric preclinical study, or review paper

11 eligible studies included

188 ineligible articles:

Pre-2000 or duplicates

13 papers excluded as insufficient ARDS specific data provided or ECCO_2_R therapy not applied for minimum timeframe

**Figure 1.** Preferred Reporting Items for Systematic Reviews and Meta-Analyses (PRISMA) flow chart of study selection for venovenous ECCO2R use in moderate to severe ARDS

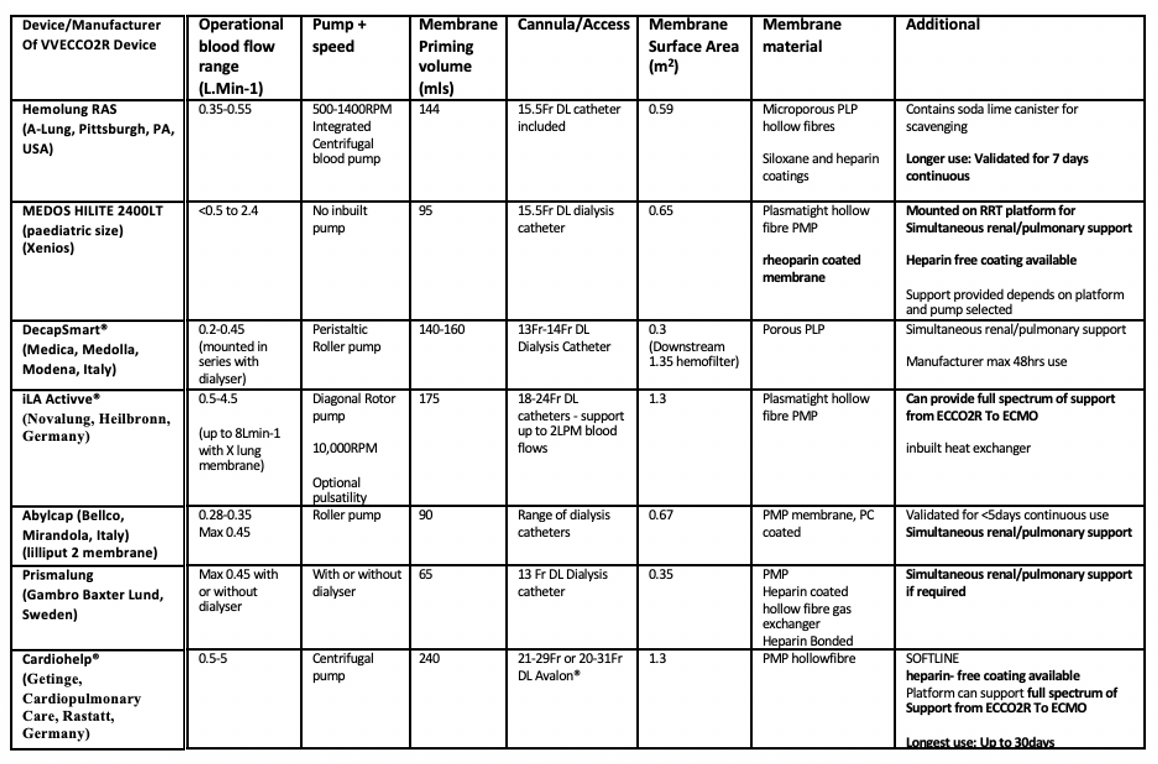

**Table 1a**: Modern Venovenous ECCO2R platforms. Device operational characteristics and special features

| Study | ECCO_2_R Device | Access | Anticoagulation | BFR (ml/min) | Membrane Area (m^2^) | CO2 removal  ( ml/min) |
| --- | --- | --- | --- | --- | --- | --- |
| McNamee 2021 | Hemolung ® RAS - A-Lung technologies | 15.5 Fr DL - IJV/FV | NR | 350-450 | 0.59 | 83.4 (34.5) |
| Goursaud 2021 | Prismalung® Gambrio-Baxter | 13-14Fr DL IJV/FV | AntiXa 0.54 (0.45;0.81) | 400 (400-400) | 0.32 | NR |
| Augy 2019 | Hemolung®RAS, iLA Activve(HF) | 15.5Fr DL RIJV/FV or 18 (IJV)-24Fr(FV) DL* | AntiXa 0.3-0.6 | ^350-500 (L) 500-1500 (H)  NR | NR | NR |
| Combes 2019 | Hemolung RAS, iLA Activve, Cardiohelp®HLS 5.0 Device* | 15.5Fr DL FV/IJV (L)OR 18-20Fr (H)* | APTT 49.1±14.9 (L)  57.6±21.6 (H)* | 440 (L)  970 (H) | 0.59 (L)  1.3 (H) | NR |
| Schmidt 2018 | Prismalung® Gambro-Baxter | 13Fr DL RIJV/FV | APTTR 1.8±0.6 | 421±42 | 0.32 | 51±26 |
| Winiszewski 2018 | Hemolung®, Prismalung®,  iLA Activve®, Cardiohelp® | NR | AntiXa 0.14-0.47 | NR | NR | NR |
| Pepperstraete 2017 | Abylcap® (Bellco) | 13.5Fr DL FV | ACT 211s (186;225) | 400 (399;410) | 0.67 | NR |
| Fanelli 2016 | Hemolung ® RAS - A-Lung technologies | 15.5 Fr DL - IJV/FV | APTTR 1.77±0.7 | 435±60 | 0.59 | 81±9 |
| Allardet-Servent 2015 | MEDOS HILITE 2400LT membrane with Prismaflex® RRT | 15.5Fr DL IJV | ACT 2±0.9 | 410±30 | 0.65 | 83±20 |
| Terragni 2009 | Decap® Hemodec RRT | 14Fr DL or 2x 8Fr SL FV | APTTR 1.1-1.7 | 357±75 | 0.33 | NR |

**Table 1b.** ECCO2R device, access and anticoagulation technical data by study

*SUPERNOVA Trial and Augy et al. 2019 both utilised both high and low extraction vvECCO_2_R devices:

larger venous access reserved for Higher blood flows.

^NB: Measured blood flow rates *not reported* but authors described typical flows for devices used

H= High flow	L= Low flow IJV = Internal jugular vein	FV= femoral vein DL= double lumen	SL = single lumen

NR= not reported ACT = activated clotting time APTT(R)= activated partial thromboplastin time (ratio)

| **Study** | **Age (Yrs)** | **PaO_2_:FiO_2_**  **(mmHg)** | **Reported Mortality** | **ICU LOS (days)** | **Duration therapy** | **Time: ventilation to ECCO2R** | **SOFA Score at baseline** | **SAPSII at baseline** | **RRT utilisation** |
| --- | --- | --- | --- | --- | --- | --- | --- | --- | --- |
| **McNamee 2021 ^*** | 60.2 (50.6-69.0) | 118.1 (96.0-134.3) | 38% 28d | 14 (7-26) | 4(2) d | NR | 10 (7-12) | NR | NR |
| **Goursaud 2021** | 64 (57;76) | 108.5 (96.5;136.3) | 44% 28d | 15 (11-20) | >24hrs | 2(1-3.8) | 6 (5-8) | 42 (39-45) | 17% |
| **Combes et al**  **2019** | 60.2±14.0 | 101.2±34.5 | 27% 28d | NR | 5(3-8) d | NR | 7.42±3.22 | 45.9 (±15.5) | NR |
| **Augy 2019** | 66 (63-77) | 131 (100-190) | 71%^§^ | NR | 4 (2-6) d | NR | NR | 48(43-62) | 11% |
| **Schmidt**  **2018** | 60±12 | 188±75 | 15% 28d | 18 (14-41) | 31±22 hrs | 4 (2-7)d | 9.3±4.3 | 56 (±21) | NR |
| **Winisziewski**  **2018** | 63 (59-68)* | 145 (116-161) | 31% 28d | 18 (11-26) | 6 (5-9) d | 3 (1-5)d | 10 (7-12) * | NR | 21%* |
| **Peperstraete**  **2017** | 50.5 (34.8;63.3) | 83 (67.6;121.1) | 40% 28d | NR | 6 (5;12) days | 26  (20.5-67.5) hrs | NR | 69.5 (57.75-79.25) SAPS 3 | 40% |
| **Fanelli 2016** | 55±19 | 159 ±34 | 47% 28d | NR | >24hrs | NR | 10±4 | 51±14 | 0% |
| **Allardet-Servent 2015** | 70±9 | 133±41 | 82% ICU | 40 (12-66) | NR | 2(1-4)d | 14±4 | 69±13 | 100% |
| **Terragni 2009** | 65.8±12.22 | 147±56 | NR | NR | 141±69 hrs | NR | NR | 48 (±20) | NR |

**Table 2a.** Demography and outcome data

Data expressed as Mean (±SD) or as Median (IQR)

^* Intervention group

§ Timeframe unspecified

*data here expressed as recorded for the whole cohort, not specifically ARDS patients

Table 2b.

**Demographics**

Male (9 studies)

- 68.4% (95%-CI: 63.5% to 73.1%), i^2^ = 0.0%

Age (10 studies)

- 62.2 years (95%-CI: 59.9 to 64.6), I^2^ = 61.3%

BMI (6 studies)

- 28.0 (95%-CI: 26.2 to 29.8), I^2^ = 59.1%

Baseline PF ratio (10 studies)

- 141.03 (95%-CI: 122.41 to 159.66), I^2^= 90.9%

Mortality (10 studies)

- 41.6% (95%-CI: 30.0% to 53.5%), I^2^ = 73.5

**Meta-regression Analysis**

| Covariate | Study | Estimate | LCI | UCI | P |
| --- | --- | --- | --- | --- | --- |
| **Age** | **7** | **0.1285** | **0.0207** | **0.2363** | **0.020** |
| Baseline CO2 | 7 | 0.0029 | -0.0781 | 0.0839 | 0.94 |
| Baseline DP | 7 | -0.1529 | -0.4047 | 0.0990 | 0.23 |
| Baseline PF | 7 | -0.0122 | -0.0266 | 0.0022 | 0.096 |

**Table 3.** Meta-regression Analysis with respect to the Primary outcome (change in Driving Pressure)

LCI=Lower Confidence interval, UCI= Upper Confidence interval

+ve = smaller reduction in DP

-ve = larger reduction in DP

| **Study** | **Bleeding** | **Transfusion: PRC (units)** | **Platelet Transfusion** | **Anticoagulation** |
| --- | --- | --- | --- | --- |
| **McNamee 2021** | Total patient bleeding events = 33  ICH = 9 SAEs (4.5%) – 5 considered likely due to ECCO2R  Other site bleeding – 18 in intervention group  4 considered due to ECCO2R (2%) | NR | NR | Heparin: target aPTTR 1.5-2 |
| **Goursaud 2021** | 1 ICH reported (5.6%) | NR | NR | AntiXa 0.54 (0.45-0.81) IU/ml |
| **Combes 2019** | 13 pts (14%)  6% (requiring Transfusion) | NR | NR | aPTT 35-80 |
| **Augy 2019** | Total = 17 pts (24.3%)    16 (30.2%) Hemolung®  1 (5.9%) iLa Activve®  Bleeding cause of discontinuation in 6 pts | 7 pts received transfusion  3 pts - catheter embolisations  1 pt: surgical management |  | AntiXa 0.3-0.6 IU/ml |
| **Schmidt 2018** | 2 pts (10%) – none required transfusion | 0 | NR | aPTTR 1.8±0.6 |
| **Winiszewski 2018^¶^** | Total 16 pts (48%)  By device:  9 (53%) Hemolung®  4 (40%) Prismalung®  3 (75%) iLa Activve® | NR | 4 (12%) received | AntiXa 0.22-0.35 IU/ml |
| **Peperstraete 2017** | BARC and GUSTO definitions used*  4 pts(40%) moderate/3a  1 pts(10%) mild/2 | 5 pts (50%) received transfusion  Total units = 18  1.5 [0;3.25] | NR | D1 ACT 211s [186;225] |
| **Fanelli 2016** | 7 pts (46.7%) | D1: 2.25±0.5 (4 pts)  D2: 1.5±0.7 (2 pts)  D3: 2 (1 pt) | D1: 1.25±2.5 (4 pts) | D1 APTTR 1.77 |
| **Allardet-Servent 2015** | NR | NR | NR | ACT 2±0.9 during study |
| **Terragni 2009** | No significant bleeding | 0 | NR | Max aPPTR 1.5±0.2 |

**Table 4.** Bleeding and Anticoagulation

SAE – significant adverse event ¶ Bleeding reported for whole cohort (11 COPD pts and 16 ARDS patients)

*GUSTO and BARC: “Bleeding Academic Research Consortium Definition for Bleeding” and the “Global Utilization of Streptokinase and Tpa for Occluded arteries definition of bleeding”

ICH=intracerebral haemorrhage, aPTT/R= (activated partial thromboplastin time/ratio), ACT= activated clotting time

Mean complication rates calculated across trials reporting the complication to minimise bias from under-reporting.

***=**Used GUSTO (Global Utilization Of Streptokinase And Tpa For Occluded Arteries) criteria**;** ‘ = median PRC transfusion requirement **;** ^= Biological haemolysis/Clinically significant haemolysis (biological= serum free Hb >100mg/l); Total Cohort complication rates reported

(ie. ARDS+non-ARDS patients: Winiszewski 2018 + Augy 2019
